# NMR metabolomic modelling of age and lifespan: a multi-cohort analysis

**DOI:** 10.1101/2023.11.07.23298200

**Authors:** Chung-Ho E. Lau, Maria Manou, Georgios Markozannes, Mika Ala-Korpela, Yoav Ben- Shlomo, Nish Chaturvedi, Jorgen Engmann, Aleksandra Gentry-Maharaj, Karl-Heinz Herzig, Aroon Hingorani, Marjo-Riitta Järvelin, Mika Kähönen, Mika Kivimäki, Terho Lehtimäki, Saara Marttila, Usha Menon, Patricia B. Munroe, Saranya Palaniswamy, Rui Providencia, Olli Raitakari, Floriaan Schmidt, Sylvain Sebert, Andrew Wong, Paolo Vineis, Ioanna Tzoulaki, Oliver Robinson

## Abstract

Metabolomic age models have been proposed for the study of biological aging, however they have not been widely validated. We aimed to assess the performance of newly developed and existing nuclear magnetic resonance spectroscopy (NMR) metabolomic age models for prediction of chronological age (CA), mortality, and age-related disease.

98 metabolic variables were measured in blood from nine UK and Finnish cohort studies (N ≈ 31,000 individuals, age range 24-86 years). We used non-linear and penalised regression to model CA and time to all-cause mortality. We examined associations of four new and two previously published metabolomic age models, with ageing risk factors and phenotypes. Within the UK Biobank (N≈ 102,000), we tested prediction of CA, incident disease (cardiovascular disease (CVD), type-2 diabetes mellitus, cancer, dementia, chronic obstructive pulmonary disease) and all-cause mortality.

Cross-validated Pearson’s *r* between metabolomic age models and CA ranged between 0.47–0.65 in the training set (mean absolute error: 8-9 years). Metabolomic age models, adjusted for CA, were associated with C-reactive protein, and inversely associated with glomerular filtration rate. Positively associated risk factors included obesity, diabetes, smoking, and physical inactivity. In UK Biobank, correlations of metabolomic age with chronological age were modest (*r* = 0.29–0.33), yet all metabolomic model scores predicted mortality (hazard ratios of 1.01 to 1.06 / metabolomic age year) and CVD, after adjustment for CA.

While metabolomic age models were only moderately associated with CA in an independent population, they provided additional prediction of morbidity and mortality over CA itself, suggesting their wider applicability.

## Introduction

Ageing can be broadly defined as a time-dependent decline of functional capacity and stress resistance, associated with increased risk of morbidity and mortality (Burkle et al., 2015). The rate of ageing may vary between individuals and groups due to both environmental stressors, including lifestyle and social adversity (Stringhini et al., 2018), and genetic factors (McDaid et al., 2017). This divergence in the rate of ageing can lead to discrepancies between “biological” and chronological age. Markers of biological age may allow improved prediction of health- and life-span than chronological age itself and allow identification of vulnerable individuals (Ferrucci, Levine, Kuo, & Simonsick, 2018).

Recently, high throughput ‘omic’ methods, which provide simultaneous quantification of sets of multiple molecular features, have been used to develop ‘biological clocks’ that provide a global measure of changes with age at the molecular level (Rutledge, Oh, & Wyss-Coray, 2022). Metabolomics, the global profiling of small molecules with a molecular weight of <1,500 Da in the body, has emerged as a promising analytical approach for assessing molecular changes with age at the population level (Panyard, Yu, & Snyder, 2022; Robinson & Lau, 2023). Overall, the rate of metabolism declines with age (Pontzer et al., 2021) and more specifically all ageing hallmarks are expected to have detectable effects on the metabolome, including hallmarks of cellular ageing such as nutrient sensing, mitochondrial dysfunction, and altered intracellular communication which directly relate to metabolic alterations (Lopez-Otin, Galluzzi, Freije, Madeo, & Kroemer, 2016; Nilsson et al., 2019).

We previously reported a metabolomic clock based on untargeted mass-spectrometry (Robinson et al., 2020) in a cohort of around 2,000 people, observing strong prediction of chronological age (CA) in internal test sets, associations between metabolomic age and non-communicable disease risk factors, and enrichment of known ageing pathways among model predictors. However, this clock cannot be easily applied to other datasets due the untargeted nature of the data used. An alternative approach is to use nuclear magnetic resonance spectroscopy (NMR), a metabolomic platform that provides more precise quantification enabling more straight-forward application across studies. Van den Akker et al. (van den Akker et al., 2020) used the Nightingale platform of NMR-based metabolomics in blood, to linearly model CA in a large Dutch Biobank sample of 25,000 people from 26 cohorts (age range 18-85), finding their metabolomic age measure was predictive of cardiovascular events and mortality. While their metabolomic age measure was strongly correlated with CA in an internal test set, internal validation may provide overoptimistic assessments of model performance (Rodriguez-Perez, Fernandez, & Marco, 2018) and their measure remains to be widely tested in external datasets.

When developing biological age clocks, two divergent approaches have emerged: training on chronological age, which will identify molecular features and pathways that change with CA but may be less sensitive for assessing age-related health status; and training on lifespan (i.e. time-to-all-cause mortality) which may more accurately reflect one’s age-related health status, yet will also assess early-effects of disease in addition to intrinsic biological ageing mechanisms (Bernabeu et al., 2023). In this regard, the multivariable NMR-based metabolite score of all-cause mortality developed by Deelen *et al*. in 44,000 people may be considered a biological age marker as it explicitly assesses remaining lifespan. The model was found to have greater predictive accuracy than a model containing conventional risk factors (Deelen et al., 2019).

In the present study, we aimed to develop new NMR-based metabolomic ageing models, incorporating variable selection, non-linear modelling, and lifespan information, within nine UK and Finnish cohorts of 38,000 samples covering most of adult life. To judge their potential utility for the assessment of differential metabolic ageing, we assessed and compared their associations with ageing risk factors, phenotypes, and cardiovascular disease and mortality incidence. Finally, to understand the reproducibility of the NMR-based metabolomic ageing models, we tested their performance, alongside the previously published Akker *et al*. and Deelen *et al*. models, in the UK Biobank (UKB, N = 102,000 individuals) for the prediction of CA, mortality, and a diverse range of age-related diseases.

## Methods

### Study population

The study included six British cohorts participating in the UCL-LSHTM-Edinburgh-Bristol (UCLEB) Consortium (Shah et al., 2013): the MRC National Survey of Health and Development (NSHD), the Caerphilly Prospective Study (CAPS), the British Women’s Heart and Health Study (BWHHS), the Southhall and Brent Revisited Study (SABRE), the Whitehall-II study (WHII) and the UK Collaborative Trial of Ovarian Cancer Screening Longitudinal Women’s Cohort (UKCTOCS). Two studies from Finland were included: the 1966 Northern Finland Birth Cohort (NFBC1966) and the Young Finns Study (YFS). In addition, we included the British Avon Longitudinal Study of Parents and Children (Boyd et al., 2013), which included samples from fathers (ALSPAC-partners) (Northstone, 2023) and mothers (ALSPAC-mothers) (Fraser et al., 2013). ALSPAC-partners and ALSPAC-mothers were considered as different cohorts and analyzed separately. Longitudinal samples were available from SABRE at 2 timepoints (SABRE1, SABRE2) which were collected between 1988–91 and 2008–11. Follow-up samples were available from NFBC1966 at 2 timepoints when participants were 31 [NFBC1966 (31y)] and 46 years old [NFBC1966 (46y)], and longitudinal YFS samples available were followed-up in 2001 (YFS2001), 2007 (YFS2007) and 2011 (YFS2011). These follow up samples were analyzed separately since follow-up clinics and sampling were conducted on separate occasions. UKB study samples (N ≈ 102,000) were used for model validation in this study. Ethical approval for each cohort study was obtained from the Local Research Ethics Committees. Informed consent for the use of data collected via questionnaires and clinics and analysis of biological samples was obtained from all participants. Additionally, the current study was approved by the Imperial College Research Ethics Committee (Reference: 19IC5567). Details on individual cohort characteristics are listed in Supplementary Table 1.

### Metabolomic data acquisition and pre-processing

A high throughput ^1^H NMR spectroscopy metabolomics platform (Brainshake Ltd./Nightingale Health©, Helsinki, Finland) was applied to fasted blood serum or plasma samples. The assay provides concentration measurements for a range of metabolite variables including lipoprotein subclasses and individual lipids, fatty acids, glucose and various glycolysis precursors, ketone bodies and amino acids. The NMR platform also reports on average sizes of lipoprotein particle subclasses VLDL, LDL, and HDL. Details of this platform have been published previously (Soininen et al., 2009; Wurtz et al., 2017). Two hundred and thirty-three lipid and metabolite measures were initially obtained from the assay platform, although some of the metabolic measures were frequently missing in one or more cohorts and were removed from subsequent analysis. Acetoacetate, pyruvate, glycerol, glycine, diacylglycerol, conjugated linoleic acid, and estimated description of fatty acid chain length were excluded as a result. Derived metabolic variables, including variables expressed as ratios or percentages were also excluded to limit data redundancy. Additionally, we also examined correlations of metabolic variable signals derived from Nightingale pre-2020 and post-2020 quantification protocols available for 6,446 YFS samples and excluded variables with Pearson’s *r* < 0.7. The remaining 98 well quantified metabolic variable signals were considered in the main part of this study (Supplemental Data Table 2). Multivariate outlier detection was carried out per study cohort, using the *pcout* function from R package *mvoutlier*, to remove sample outliers from the study. The method is based on principal components analysis and observations were considered location outliers if they have been assigned a weight ≤ 0.1, and these were subsequently removed from the study. Six thousand, one hundred four samples were removed as a result, and the number of samples after quality control was 37,888.

Additionally, to minimize bias originated from pre-analytical and analytical differences in amongst the study cohort datasets, we calibrated the metabolic data between cohorts and visits using methodology as described in Makinen et al. (Makinen et al., 2022) as part of the data pre-processing. Whitehall II (WHII) was a mixed-sex cohort in the middle of the age range in amongst our samples and was thus defined as the reference dataset in the calibration, with all other study cohort datasets were normalized against the WHII samples. During the cohort data calibration, a subset of samples of matching demographic characteristics, including age, sex, body mass index (BMI), and ethnicity were selected from both the target and the reference datasets, and scaling factors were then estimated per metabolic variable and subsequently applied to the full cohort data in the target sets. The distribution of values in the 98 metabolic variables were broadly normal, as determined through visual inspection and thus no other data transformation procedures were applied.

### Metabolome wide association study (MWAS) of age and mortality

To understand individual metabolite associations with ageing, we first performed univariate analyses of metabolic variables with age and mortality. Cohort-stratified metabolome wide association study (MWAS) of age were assessed using multiple linear regression adjusted for sex, BMI, and ethnicity. Age stratified MWAS of age were performed to examine the consistency of age-metabolite associations across the life-course, and these analyses were additionally adjusted for cohort. The following age group strata were used: 20-35, 35-40, 40-45, 45-50, 50-55, 55-60, 60-65, 65-70, and > 70. Multiple Cox proportional hazard regressions (*survival* R package) adjusted for chronological age, sex and BMI were used to estimate the associations with mortality, within the UKCTOCS, WHII and SABRE cohorts where this information was available. Inverse variance-weighted fixed effect meta-analyses were used to pool study cohort estimates, Benjamini & Hochberg’s false discovery rate (FDR) was used when accounting for multiple testing, with an FDR-corrected *q* < 0.05 denoting significance, and heterogeneity amongst the cohorts/ age group strata was assessed using Cochran’s Q test and I^2^ using the *meta* R package. Variables with I^2^ values > 0.75 were considered of high heterogeneity, whilst those with I^2^ values < 0.25 were considered of low heterogeneity.

### Multivariable predictive modelling of ageing

NSHD and NFBC1966 were birth cohorts and were excluded from model training since study participants all share identical CA and would therefore likely bias the training sample set. Consequently, the training sample set consisted of 26,640 samples from eight study cohorts. To avoid problems associated with multi-collinearity in model training and improve model stability, a pruned variable set was generated from the full set of predictors using sequential backward stepwise selections and the variable inflation factor (VIF), derived using *vif* function in the *car* R package, as selection criteria. Starting with all 98 predictors as model inputs, in a stepwise fashion, the variable with the largest VIF value was removed and a new model was generated with one less variable than in the previous step, until no variables had a VIF ≥ 5. This yielded 24 variables (Supplemental Data Table 2), which were then used as input predictors in our multivariable chronological ageing models. Seven-fold cross validation and leave-one-cohort out (LOCO) validation were used to assess model stability and prediction performance during training.

#### Elastic Net and MARS models

Multivariable models of study CA were constructed using elastic net regression (*glmnet* and *caret* R packages), and 2^nd^ degree multivariate adaptive regression splines (MARS, *earth* and *caret* R packages) models. Elastic net is a versatile penalized linear regression model which simultaneously performs variable selection and modelling fitting. It is computationally efficient, suitable for highly correlated datasets, and resultant models are easily interpretable (Zou & Hastie, 2005). The alpha parameter in *glmnet* was pre-selected as 0.5 in the elastic net model. The MARS approach is suitable for regression problems when the relationship between predictors and response variables are non-linear, as the model takes the form of an expansion in product spline basis functions (Friedman, 1991). A second-degree MARS model was used as it is suitable for modeling quadratic predictor-response relationships and is considered efficient when the number of model predictor variables is relatively small. Model variable importance scores (VIP) were evaluated in the training sample data using the *vip* R package.

#### Study mortality score

Instead of training the metabolic data on chronological age, multivariable modelling was performed with survival treated as dependent variable in a penalised Cox regression. Study samples from WHII, SABRE, and UKCTOCS were used for model training, which was performed using the *glmnet* R package with alpha parameter in the elastic net model selected as 0.5. The 24 metabolite variables, and covariates comprised of age, sex, BMI, ethnicity, and cohort were included as model input predictors. After including only metabolic variable predictors and excluding contributions from other covariates, the resultant model was considered as the study mortality score, and these were subsequently scaled to the means and standard deviations of CA in the study cohort sample data to render score units in years.

#### Phenotypic ageing

The phenotypic ageing model represented a hybrid approach, and it simultaneously incorporated metabolic information of both age and mortality into the model training process. Whilst this model was trained on sample chronological age, we additionally introduced a weighted approach to allow differential predictor shrinkage based on the direction and strength of their associations with mortality in our study samples. More specifically, the differential weights on the predictors were introduced as penalty factors into the *glmnet* model. Whereas a penalty factor of 0 would suggest no shrinkage, metabolic variables with large penalty factors would be heavily penalized in the model. *P-*values obtained from proportional hazard regressions of metabolic variables on mortality were applied as model penalty factors, and in addition, variables showing opposing direction of associations with age and mortality were assigned a penalty factor of 1. This approach has the effect of enhancing the influence of metabolic variables that are closely associated with mortality/ health outcome whilst still providing a direct prediction on sample chronological age.

#### Akker et al. and Deelen et al. models

The Akker *et al*. model predicts CA (in years) directly. Model weight/ coefficients were extracted from their original publication (van den Akker et al., 2020). The Deelen *et.al* model was computed using 14 log-transformed and cohort-scaled biomarkers multiplied by their weight based on *log*-hazard ratios from meta-analyses as reported in Deelen *et.al* ’s publication (Deelen et al., 2019), and subsequently summed. The resulting score was scaled to the means and standard deviations of cohort CA in the study cohort sample data to render score units in years. Acetoacetate concentrations were missing in the ALSPAC-partners and CAPS study, and these values were imputed using k-nearest neighbours method from the *impute* R package for the purpose of generating the Deelen *et al*. and Akker *et al*. model scores. The Akker *et al*. model was not applied to the UKB as two of the specified model variables have since been discontinued and were not available in the UKB dataset (Bizzarri et al., 2023).

Further details of the multivariable ageing models are provided in the supplementary material.

### Covariate coding of disease risk factors and adverse health outcomes

Hypertension was defined by doctor diagnosis in the YFS and UKCTOCS cohorts, by systolic blood pressure ≥ 140 mm Hg or doctor diagnosis in the NFBC1966, ALSPAC-mothers, ALSPAC-partners, SABRE cohorts, by use of antihypertensive medication or systolic blood pressure ≥ 140 mm Hg in the NSHD and BWHHS cohorts and by systolic blood pressure ≥ 140 mm Hg only in WHII. Diabetes was defined by doctor diagnosis in the YFS, ALSPAC-mothers, ALSPAC-partners and UKCTOCS cohorts, by glucose ≥ 7 mmol/L or doctor diagnosis in the NFBC1966, NSHD and CAPS cohorts, by fasting glucose ≥ 7 mmol/L, 2-hour postload glucose ≥ 11.1 mmol/L or doctor diagnosis in the WHII cohort, by glycated hemoglobin (HbA1c) ≥ 6.6% or doctor diagnosis in the BWHHS cohort and by fasting glucose ≥ 7 mmol/L only in the SABRE cohort. Physical inactivity was defined as no or less than once per week of moderate/vigorous physical activity in most cohorts. For CAPS and SABRE, it was defined as the lowest tertile of calculated weekly physical activity estimates. Smoking was classified as never/former versus current smoker. Alcohol consumption was defined as no/ moderate versus heavy consumption. Heavy alcohol use was defined in the NFBC1966, YFS, NSHD, WHII, CAPS, UKCTOCS and BWHHS for men as > 21 units of alcohol per week and for women as >14 units of alcohol per week. In ALSPAC-mothers and ALSPAC-partners, heavy alcohol use was defined as more than 4 times per week. Three measures of socio-economic position (SEP) were used representing the early, mid-, and later life periods: Occupation of the participants’ fathers was classified as a manual vs non-manual. Educational level was a binary indicator when comparing those with up to secondary-level schooling only with those with further or higher education. Current or last occupation of participants was classified as manual vs non-manual.

Within the SABRE and UKCTOCS cohorts, coronary heart disease events were available: we have included both non-fatal and fatal myocardial infarction, revascularization and unstable angina events in the SABRE cohort analysis, and acute coronary syndrome, myocardial infarction, angina, and other acute and chronic ischaemic heart disease events were included in the UKCTOCS analyses. Both fatal and non-fatal stroke were included in the association analyses. All-cause mortality was available in the WHI, SABRE and UKCTOCS cohorts. Within UKB, we analyzed incident all-cause mortality and cardiovascular disease (CVD), type-2 diabetes mellitus (T2DM), chronic obstructive pulmonary disease (COPD), cancer and dementia. CVD defined as the composite of myocardial infarction (MI) cases (ST-Elevation MI and Non-ST-Elevation MI) and stroke cases (ischaemic, intracerebral haemorrhage and subarachnoid haemorrhage).

### Analysis of metabolomic age with ageing risk factors, phenotypes, and incident health events

Diabetes, hypertension, obesity (BMI > 30), physical inactivity, current smoking status, heavy alcohol consumption, education attainment and occupation status were included in the non-communicable disease risk factor analyses, categorized as binary variables. Six ageing related biomarkers, including systolic (SBP) and diastolic blood pressure (DBP), pulse pressure, C-reactive protein (CRP), estimated glomerular filtration rate (eGFR), and forced expiratory volume in first second (FEV1) were available from multiple, but not all cohorts. Biomarkers were univariate scaled to facilitate cross-comparison. Summary of biological ageing markers data by cohort are shown in **Supplemental Table 1**. For the analysis of associations between metabolomic ageing models, ageing biomarkers, and disease risk factors, linear regression models were adjusted for chronological age, sex and ethnicity. To avoid including repeated samples from the same individuals from multiple follow-up visits, samples from YFS2001, YFS2007, NBFC1966 (31y), and SABRE2 were excluded from the risk factors analysis. Cox proportional hazard regressions adjusted for chronological age, sex, and ethnicity were used to estimate the associations with disease and mortality incidence (*surviva*l R package), within the WHII, UKCTOCS, SABRE cohorts and the UKB. Since all analyses were adjusted for CA, estimates can be interpreted as years of additional metabolomic age relative to CA, equivalent to formulations such as “age acceleration” and “age gap” often used within the biological clock literature. A *p* value threshold of 0.001 was chosen for the reporting of statistical significance, considering multiple testing and the number of independent tests performed. All analyses were conducted in R version 4.

## Results

### Age and lifespan associations of metabolites

Analysis of metabolic ageing included 26,640 samples (aged 24-86, 60% female) from 22,828 individuals in eight cohorts, including 728 and 1,992 participants from the SABRE and YFS cohorts respectively, who were assessed in more than one follow-up. Individual cohort characteristics can be found in **Figure 1A** and **Supplemental Table 1**.

**Figure 1.**
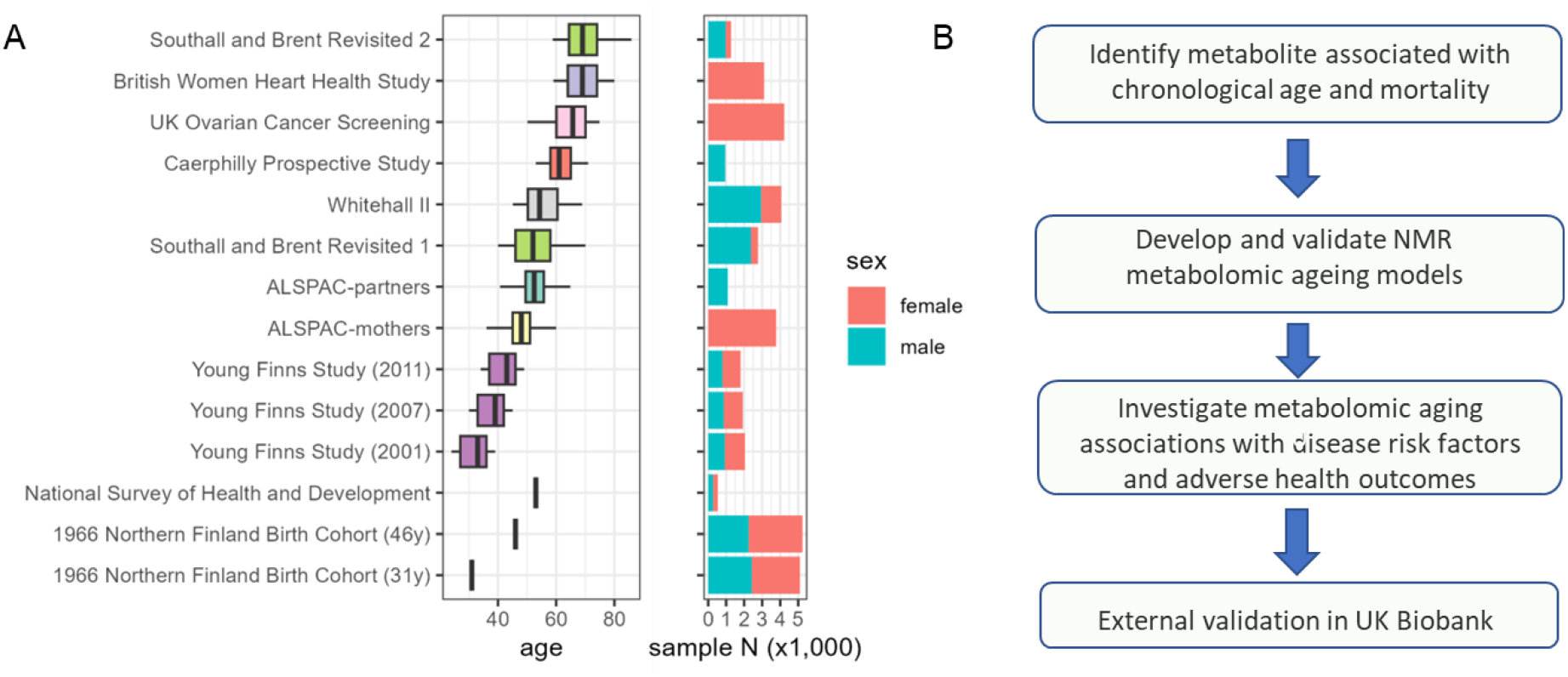
A) Study cohorts age and sex profile (37,888 samples from 30,913 subjects, of which 26,640 samples from 22,828 subjects were used for model training). The 1966 Northern Finland Birth Cohort (NFBC1966) and National Survey of Health and Development (NSHD) are both birth cohorts, where study participants all share identical age, and were only used for risk factor association analyses. B) Current study workflow.

In meta-analysis across individual cohorts and follow-ups, we identified large number of metabolic variables (N=89) tested to be significantly associated with CA after correcting for FDR at *q* < 0.05 (**Figure 2A**). For example, albumin, histidine (His), leucine (Leu), phospholipids in small HDL (S_HDL_PL), diameter for VLDL particles (VLDL_size) were found to decrease with CA; whilst citrate, glucose, creatinine, β-hydroxybutyrate (bOHbutyrate), docosahexaenoic acid (DHA), omega-3 fatty acids, glutamine (Gln), tyrosine (Tyr), phenylalanine (Phe), total free cholesterol (Total_FC) and sphingomyelins were amongst those found to increase with CA the most. (**Figure 2A, Supplemental Table 3**).

**Figure 2.**
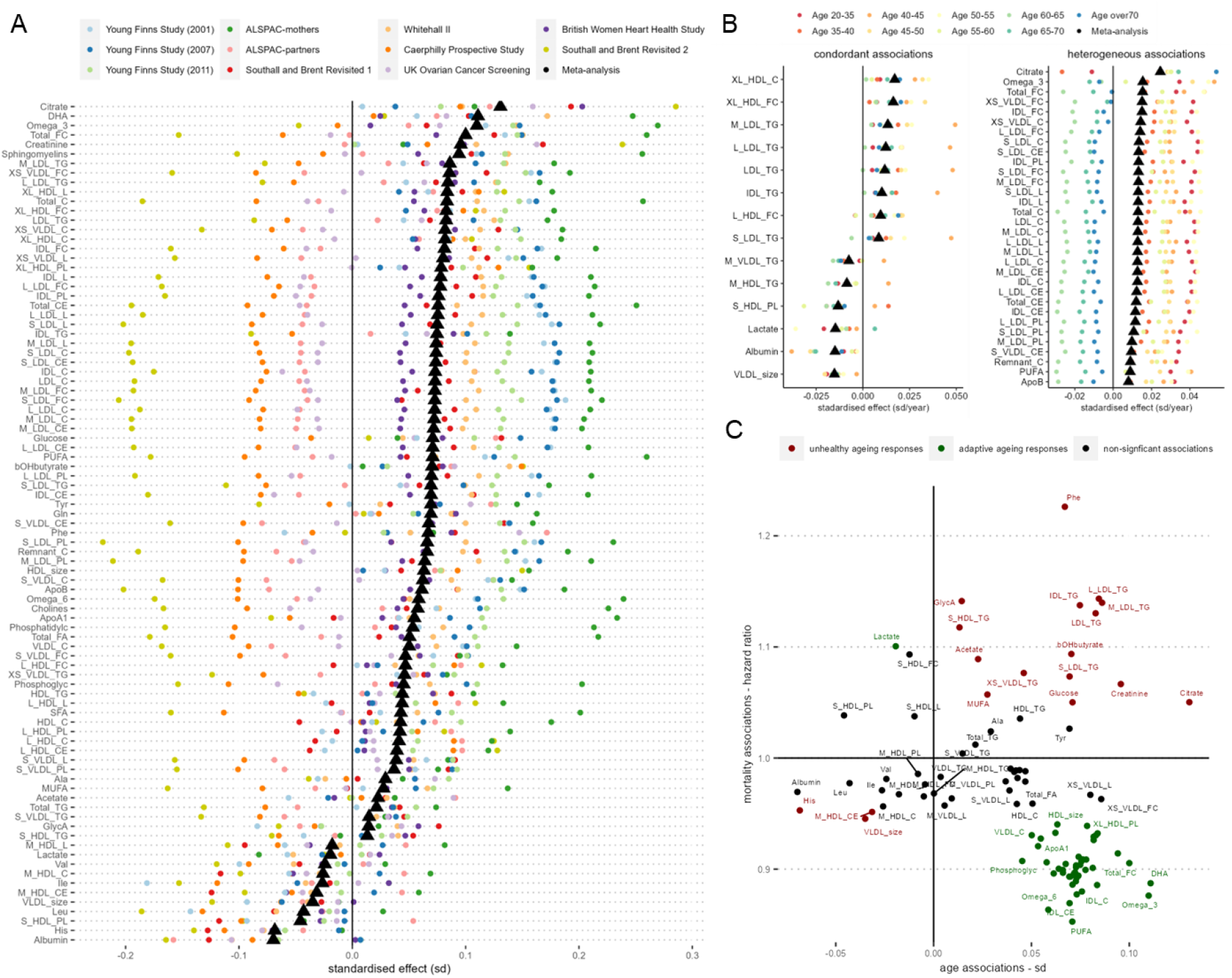
A) Age associations with NMR metabolome by individual cohort studies. Linear regression models were adjusted for sex, BMI, and ethnicity. Metabolite variables shown were found significant after FDR correction in inverse variance-weighted fixed effect meta-analyses. B) Meta-analysis of age-group stratified age-NMR metabolome associations. Linear regression models were first performed in the following age group strata: 20-35, 35-40, 40-45, 45-50, 50-55, 55-60, 60-65, 65-70, and > 70. and models were adjusted for sex, BMI, ethnicity, and cohort. Inverse variance-weighted fixed-effect meta-analysis were then performed to pool the stratified age-group model estimates. Metabolic variables found significant with FDR q < 0.05 with I^2^ values > 0.75 (high heterogeneity), or < 0.25 (low heterogeneity) were shown. C) Scatter plot of model regression coefficients of chronological age against mortality pooled hazard ratios. For mortality analysis, cohort-specific Cox proportional hazards regression models were adjusted for age, sex, BMI and ethnicity; fixed effected meta-analysis was performed to pool together individual cohort estimates. Significant metabolic associations against both age and mortality after correcting for FDR (q < 0.05) were highlighted according to whether they shared the same direction of associations: red (same direction) or green (opposing direction).The list of long names of the abbreviated metabolic variables can be found in Supplementary Table 2.

To test for consistency in response between CA and metabolic variables, we performed additional metabolome-wide association studies stratifying by age groups, additionally adjusting for cohort (***Figure 2*B)**. Metabolic variables showing consistent and positive associations with CA across age groups included triglycerides (TG) in IDL, TG variables in four LDL subfractions, and cholesterols in very large HDL particles (XL_HDL_C, XL_HDL_FC). Conversely, VLDL_size, albumin, and lactate were found to be consistently and negatively associated with chronological age. Although positively associated with CA through meta-analyses, citrate, omega-3, polyunsaturated fatty acids (PUFA), Apolipoprotein B (ApoB), and many cholesterols/ cholesterol esters and lipoprotein subfraction measurements showed heterogenous associations with CA across different age ranges. Whereas the increase in citrate levels with age appeared to be driven by older populations, the increases in many cholesterols/ cholesterol esters and lipoprotein subfraction measurements appeared to be more prominent in those aged < 60 years (***Figure 2*B, Supplemental Table 4**).

As lifespan may be considered the most relevant phenotypic endpoint for studying ageing, we examined metabolite associations with time to all-cause mortality in three cohorts (UKCTOCS, SABRE and WHII) in which mortality data were available, consisting of 10,648 individuals of whom 2,312 died during subsequent follow-up. Cohort-specific Cox proportional hazards regression models were adjusted for age, sex, BMI and ethnicity, and fixed-effect meta-analysis was performed to pool together individual cohort effect estimates (**Supplemental Figure 1, Figure 2C**). Seventeen metabolic markers were found positively associated with all-cause mortality after adjusting for false discovery rate (*q* < 0.05), which include Phe, glycoprotein acetyls (GlycA), lactate, bOHbutyrate, acetate, creatinine, glucose, monounsaturated fatty acids (MUFA), triglycerides in 7 different lipoprotein subfractions, and free cholesterol in small HDL. Forty-nine metabolic biomarkers were negatively associated with all-cause mortality, and PUFA, omega-6, omega-3 fatty acids, and cholesterols and cholesterol esters in IDL were found to be most negatively associated with mortality in our study sample (**Supplemental Figure 2, Supplemental Table 5**). Our biomarker-mortality associations study results are in good agreement with results reported by Deelen *et al*. (Deelen et al., 2019), with coefficients of mortality associations of individual metabolites of our analysis strongly correlated to the results reported (**Supplemental Figure 2**). Next, we examined correspondence between metabolites associated with age, and those associated with mortality in our dataset (***Figure 2*C**), and observed that while some age-related metabolic changes (e.g. creatinine, Phe and TG) contribute to mortality risk, at least some metabolites positively associated with CA may in fact be offering a protective effect against premature mortality (e.g. PUFA, omega-3/ omega-6 fatty acids, DHA, and cholesterol esters in IDL)

### Multivariable modelling of metabolomic ageing

Multivariable predictors for CA were trained using machine learning approaches including elastic net regression and MARS, using 24 of the most reliable and independent metabolic variables (**Figure 3A)**. Additionally, we also trained a modified elastic net model on chronological age, which we refer to as ‘phenotypical ageing’, by specifying differential model input weights based on their directionality and strength of their associations with mortality in our study samples. These three models were evaluated using 7-fold cross validations (CV) and leave-one-cohort-out validations (LOCO). Albumin and citrate were estimated to be amongst the most important predictors in all three CA models (**Figure 3B**). The overall Pearson’s correlation coefficients (*r*) between CA and the CV predicted age were 0.57, 0.65, and 0.47, and the correlations (*r*) with the LOCO predictions were 0.38, 0.37 and 0.23 respectively for the elastic net, MARS, and phenotypic age models. (**Supplemental Figure 3)**. The published Akker *et al*. chronological ageing model performed relatively poorly in our study data, giving a Pearson’s *r* = 0.26 with CA and a mean absolute error (MAE) of around 18 years of age. Among the SABRE, NFBC1966, and YFS cohorts that included repeat metabolomic data, we compared change in predicted age (δ predicted age) with change in CA (δ CA, i.e., years between assessments). We observed significant positive correlation between δ predicted age and δ CA in SABRE for all metabolomic age measures, except the Akker *et al*. model (**Figure 3C**) and general increases in median metabolomic age between follow-ups for NFBC1966 **(Figure 3D**) and YFS (**Figure 3E**). However, the models generally underpredicted δ metabolomic age relative to δ CA.

**Figure 3.**
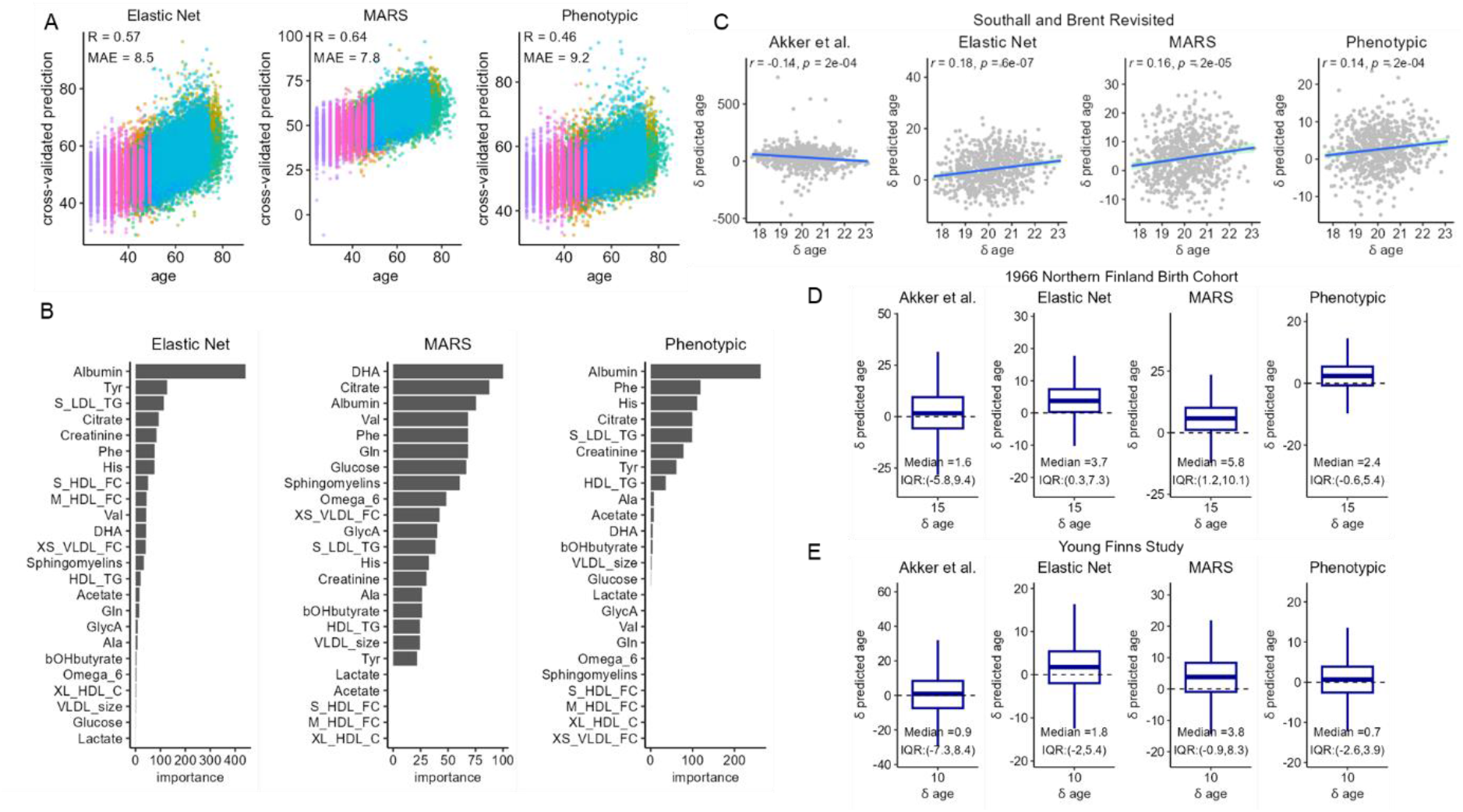
A) Scatter plots of 7-fold cross-validated predicted age against chronological age. Samples were coloured by cohorts. B) Variable importance (VIP) scores were estimated in the training samples based on the relative importance of predictors in the models C) – E) Longitudinal model predictions of changes in chronological age in Southall and Brent Revisited (SABRE), (D) 1966 Northern Finland Birth Cohort (NFBC1966), (E) Young Finns Study (YFS). Changes in the predicted age was plotted against changes in chronological age at follow-up visits in SABRE, and the boxplot shows the distribution of changes in predicted age during the 15 years and 10 years intervals between follow-up visits in NFBC1966 and YFS.

### Metabolomic ageing and age-related phenotypes

Next, we assessed and compared associations of the four metabolomic ageing models (trained on CA) and two models trained directly on mortality (Deelen *et al*. model and a new study mortality score), against non-communicable disease risk factors and six common biomarkers of ageing phenotypes, in analyses adjusted for chronological age, sex, and ethnicity. Among the risk factors, diabetes, hypertension, and obesity statuses were positively associated (p < 0.001) with all metabolomic models, and physical inactivity was also positively associated with five of the six metabolomic scores examined (**Figure 4A)**. Additionally, current smoking status and indicators of lower socio-economic positions (low education attainment and manual occupation status) were also positively associated with the Akker *et al*. and Deelen *et al*. models and the study mortality score. All metabolomic models examined were found positively associated (p < 0.001) with CRP (inflammation) and negatively associated with glomerular filtration rate (kidney function). Except for the Akker et al. model, all models were positively associated with SBP and DBP, and negatively associated with forced expiratory volume (**Figure 4B**).

**Figure 4.**
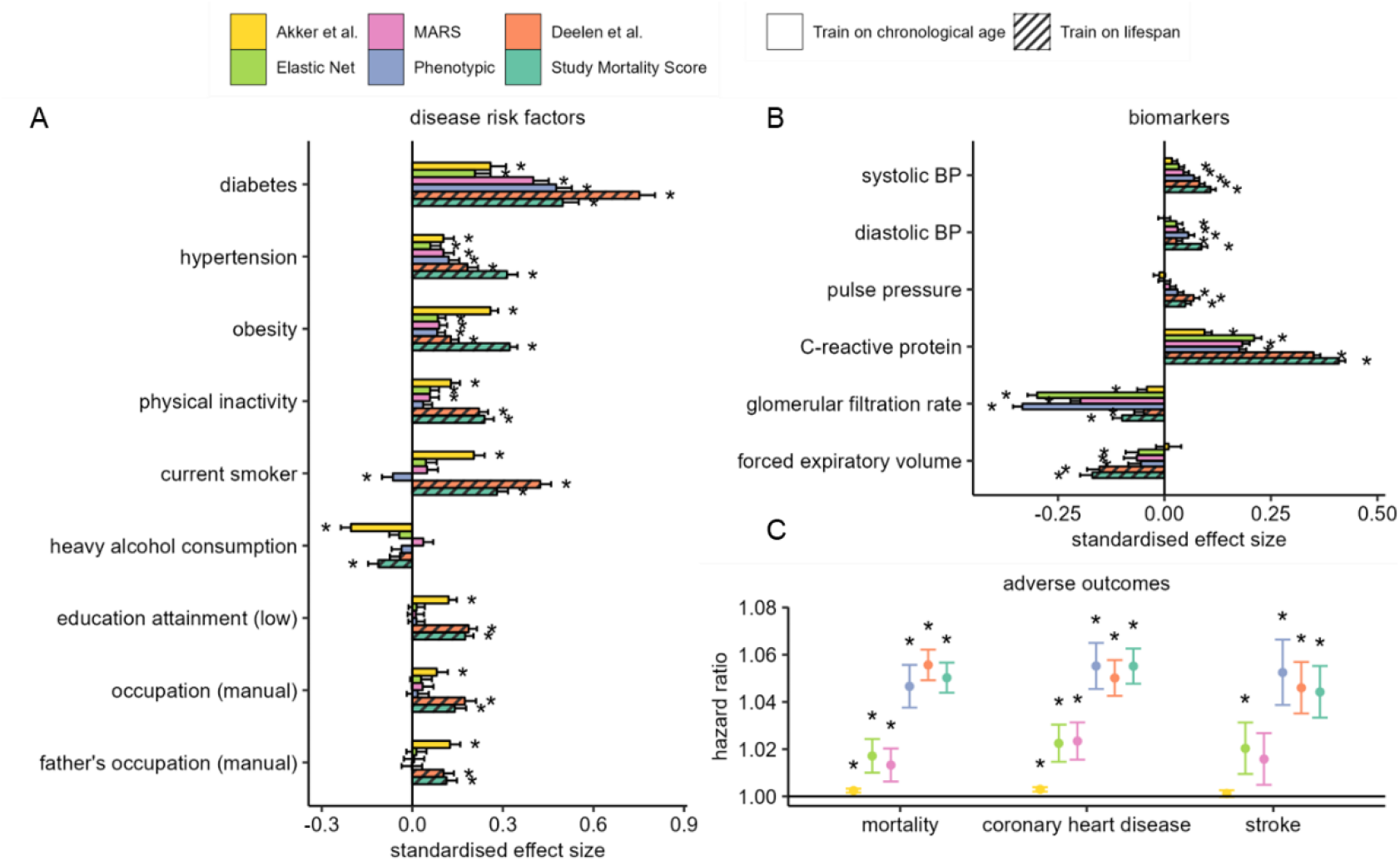
A) Associations with non-communicable disease risk factors. Estimates represent standard deviation change in metabolomic age associated with exposure which have been categorised into <u>binary</u> variables. B) Associations with age-related biomarkers. Estimates represent standard deviation (SD) change in metabolomic age associated with 1 SD unit change in biomarker levels. To avoid individuals from being accounted for more than once in the analysis, samples from YFS2001 and YFS2007, NFBC1966 (31y), and SABRE2 were excluded in the disease risk factor analysis, and subsequently up to 28,000 samples were included. C) Associations of metabolomic age models with adverse incident health events. Cox proportional regression models were adjusted for chronological age, sex, and ethnicity, and hazard ratios were estimated per unit of change in metabolomic age. 969, 638 and 715 deaths respectively in UKCTOCS, SABRE and WHII, and 1,273 and 442 coronary heart disease events, and 707 and 181 stroke events were respectively recorded in the UKCTOCS and SABRE cohorts during the subsequent follow-up period of up to 25 years. Analyses in A) – C) were based on cohort fixed effect inverse variance weighted meta-analyses and linear regression models adjusted for chronological age, sex and ethnicity. * denotes a P < 0.001 and error bars represent the lower and upper limits of the 95% confidence intervals.

Furthermore, we investigated the relationship of metabolomic age with incident health events in the cohorts with available data using Cox proportional regression models adjusted for CA, sex, and ethnicity, and fixed effect meta-analysis to combine individual cohort estimates. All metabolomic ageing models trained on CA and lifespan were significantly associated with all-cause mortality (*N _event_* = 2,312) and coronary heart disease (CHD) incidences (*N _event_* = 1,715), and with the exception of the Akker *et al*. model and MARS, all other models were significantly associated (p < 0.001) with incidence of stroke (*N _event_* = 888, **Figure 4C)** Phenotypic ageing and the two models of lifespan, Deelen *et al*. and study mortality score, were most strongly associated with adverse health outcomes, with Hazard Ratios (HRs) for all-cause mortality of 1.047 (95% Confidence Interval (CI): 1.038 – 1.056), 1.056 (95% CI: 1.049 – 1.062) and 1.05 (95% CI: 1.044 – 1.057), respectively per year of metabolomic age.

To understand the contribution of adiposity to the observed associations with metabolomic age markers, we additionally adjusted for BMI in sensitivity analysis. Associations with hypertension and blood pressure were attenuated for the chronological ageing trained model associations. However, adjusting for BMI did not significantly affect study models associations with adverse health outcomes (**Supplemental Figure 4**).

### Independent assessment in the UK Biobank

Performance of the metabolomic ageing models were tested in the large independent UK Biobank sample, comprising of metabolomic data from 101,524 individuals and accompanied by rich phenotypic and follow-up data. Models trained directly on CA provided modest predictive performance in UKB, with Pearson’s *r* with CA of 0.29, 0.33 and 0.33 for phenotypic age, elastic net, and MARS respectively. In addition, both models of lifespan (Deelen *et al*. and study mortality score) also showed significant correlations with CA (Pearson’s *r*: 0.09-0.17, p < 1 × 10^-10^) in the UKB (**Figure 5A)**.

**Figure 5.**
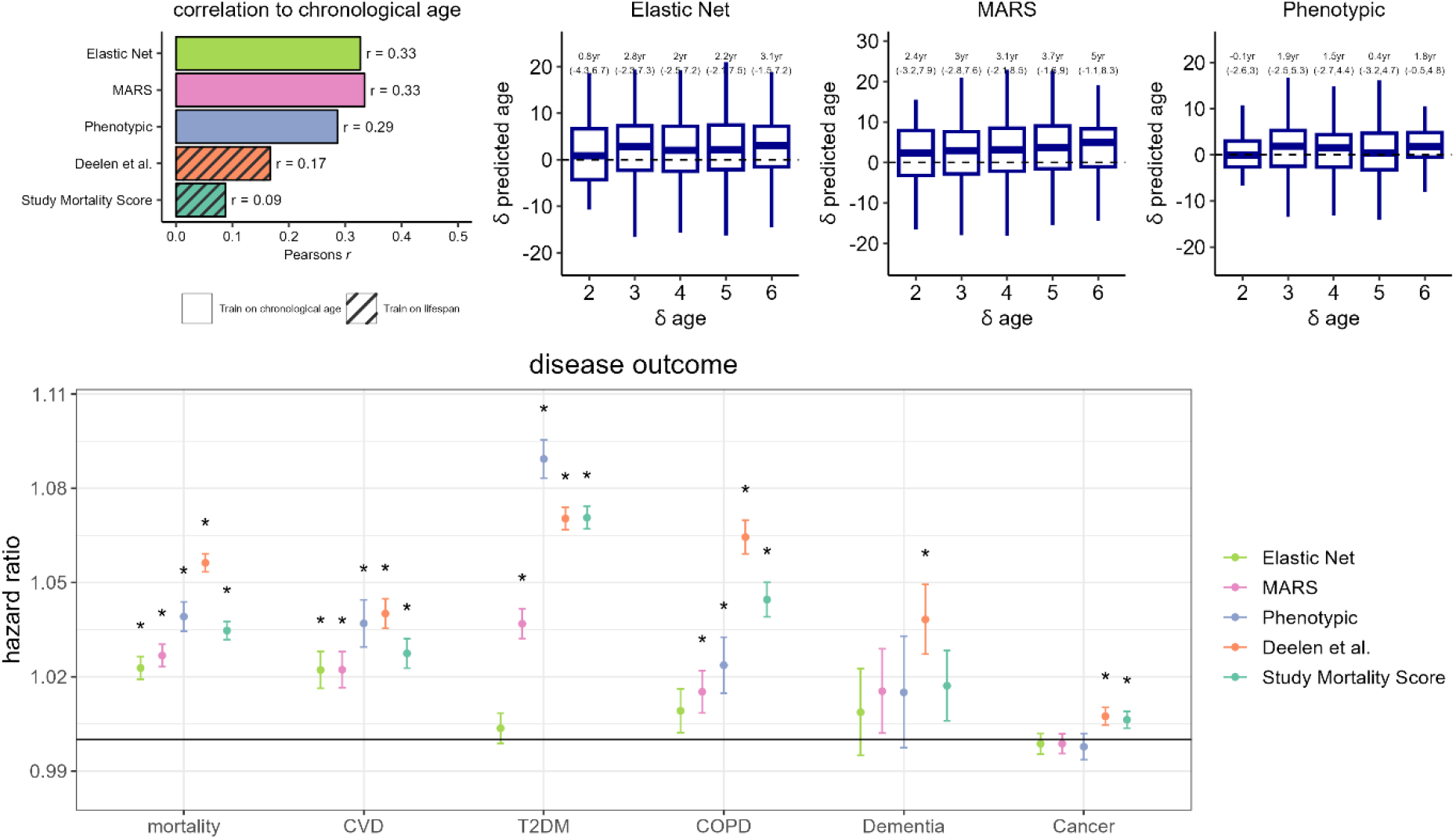
A) Assessment of metabolomic ageing model scores in UK Biobank (N _total_ = 101,524). Pearson’s correlation coefficients with chronological age are shown. B) Longitudinal assessment of metabolomic ageing model scores in UK Biobank. C) Associations of chronological age-adjusted metabolomic age scores with adverse incident events in the UK Biobank. Cox proportional regression models were adjusted for sex and chronological age. Hazard ratios were estimated based on per year of metabolomic age. * denotes a P < 0.001 and error bars represent the lower and upper limits of the 95% confidence intervals.

Among 1,108 UKB participants with longitudinal metabolomic data measured at baseline and at clinical follow-up 2-6 years later, we compared change in metabolomic age (δ metabolomic age) with change in CA (δ CA) for the CA trained models, categorized by years of follow-up. For most follow-up categories, we observed an increase in median metabolomic age over follow-up, except for phenotypic age among those only followed-up for two years. The MARS model showed the greatest concordance between δ CA and δ metabolomic age, with a median δ metabolomic age of 5 years (IQR: -1.1-8.3 year) among those with δ CA of six years (**Figure 5B)**.

Using Cox proportional regression models adjusted for CA and sex, we studied associations of metabolomic ageing models with all-cause mortality (*N _event_* = 6,645), CVD (*N _event_* = 2,585), T2DM (*N _event_* = 3,850), cancer (*N _event_* = 8,192), dementia (*N _event_* = 450) and COPD (*N _event_* = 1,814) incidences in the UKB samples (**Figure 5B, Supplementary Table 8**). All metabolomic ageing models tested were significantly associated (*p* < 0.001) with all-cause mortality and CVD. Effect estimates for all-cause mortality ranged from a HR of 1.023 per year of metabolomic age (95% CI: 1.019 -1.026) for the Elastic Net model to a HR of 1.056 (95% CI: 1.053 -1.059) for the Deelen at al. model. The next best performing model was phenotypic age (HR: 1.039 [95% CI: 1.034-1.039]), which outperformed the MARS model (HR: 1.027 [95% CI: 1.023-1.030]), which in turn outperformed the linear CA trained models for prediction of all-cause mortality. A similar pattern was observed for CVD. All models except the elastic net model were also found to be significantly associated with incidences of T2DM and COPD. The best performing model for T2DM prediction was phenotypic age (HR: 1.089 [95% CI: 1.083-1.095]) and the Deelen et al. model performed the best for prediction of COPD (HR: 1.064 [95% CI: 1.059-1.070]). Both the mortality score and the Deelen *et al*. model showed small significant association with cancer incidence, while only the Deelen et al. model was associated with dementia incidence.

## Discussion

In one of the largest epidemiological metabolomic studies to date, we have developed and tested the performance of various multivariable metrics to assess ageing as a biological process.

In brief, metabolomics data was generated through NMR spectroscopy in blood (serum or plasma) from nine UK and Finnish cohort studies, covering an age range from 24-86 years. We used multivariate adaptive regression splines (MARS) and penalised regression models to predict CA and mortality. Alongside two published metabolomic prediction scores (“Akker *et al*.” trained on chronological age, and “Deelen *et al*.”, trained on all-cause mortality), we examined associations of new chronological age-adjusted metabolomic age models with ageing phenotypes. These metabolomic measures were associated with blood pressure parameters and C-reactive protein levels and inversely associated with glomerular filtration rate. Risk factors associated with age-adjusted metabolomic age scores included obesity, diabetes, smoking, physical inactivity, and low education level. In independent testing in the UK Biobank, correlations with CA were modest, yet all metabolomic model scores predicted all-cause mortality and CVD.

### Performance of different models: prediction of chronological age

One criterion that a biological age estimator should fulfil is that should change with CA (Moskalev, 2019). When we compared our models to the UKB set, correlation with CA was more modest. The MARS model performed the best, based on model fit in the training set and associations with CA and δ CA in the UKB, indicating the value of incorporating non-linear modelling. However, taken together, models trained on CA provided only moderately improved age prediction performance compared to models trained on lifespan. The models trained on CA in our study also apparently outperformed the previously published Akker *et al*. model (albeit tested in different independent populations), despite it being trained on a similarly sized dataset. This difference may be due to the additional pre-processing and variable selection steps applied, thereby increasing model stability, and potentially due to use of fasting samples only in our training set (i.e., not in UKB) reducing the influence of recent food intake on metabolite levels. Overall, as predictors of CA across independent test sets, models based on NMR metabolomic data fall a long way short of gold-standard data types such as DNA methylation (Hannum et al., 2013; Horvath, 2013), although perform somewhat similarly to telomere length (Bekaert, De Meyer, & Van Oostveldt, 2005; Vaiserman & Krasnienkov, 2021).

### Prediction of mortality and disease incidence

Biological age estimators should also predict mortality better than CA and predict the early stages of a specific age-related disease (Ferrucci et al., 2020; Levine, 2013). To test this, we assessed the metabolomic scores adjusted for chronological age, against mortality and incidence of age-related disease. All models were able to predict mortality in both the training set and the UKB, with generally similar estimates in both populations, with the greatest effect size seen for the Deelen mortality estimator. The Deelen *et al*. model was also the only model that could predict incidence of all age-related diseases tested (CVD, T2DM, cancer, dementia, and COPD) in UKB suggesting it is able to capture generalizable age-related disease susceptibility. Phenotypic age performed well in terms of mortality and disease prediction, while still offering comparable associations with CA in UKB to the models trained purely on CA. Effects sizes in predicting time-to-death for the presented metabolic models, particularly Deelen *et al*. and phenotypic ageing model, were comparable to those of reported biological age assessments based on clinical markers, such as BioAge and PhenoAge (Kuo, Pilling, Liu, Atkins, & Levine, 2021), and epigenetic clocks such as the Horvath, Hannum, and DNAm PhenoAge clocks (Hannum et al., 2013; Horvath, 2013; Levine et al., 2018), although smaller than the GrimAge epigenetic clock (Lu et al., 2019). However, the advantage of age models based on NMR metabolomic data compared to other more complex indicators is that a single analysis is required rather than assaying multiple clinical markers and they are relatively cost-effective, especially compared to acquisition of epigenetic data.

### Physiological interpretation

In meta-analysis, we observed generally consistent decreases with age in metabolic measures including albumin, a marker of liver and kidney function, essential amino acid histidine, the branched-chain amino acid leucine, phospholipids in small HDL, and the diameter of VLDL. Conversely, increases with age were observed in citrate, glucose, amino-acids creatinine and glutamine, aromatic amino acids tyrosine and phenylalanine, the ketone body β-hydroxybutyrate, omega-3 fatty acids, the degree of unsaturation of fatty acids, triglycerides, and large and very large HDL. The increase in triglyceride levels is well-established in ageing, as it reflects changes in plasma TG clearance, adipose tissue lipolysis, and the partitioning of fat (Spitler & Davies, 2020). Citrate, in addition to its key role as an energy hub metabolite, may be released through increased bone resorption (Granchi, Baldini, Ulivieri, & Caudarella, 2019) and has also recently been demonstrated to be an independent marker of extracellular senescence in *in-vitro* models (James, Bennett, & Parkinson, 2018). Also, increased blood level of phenylalanine with age has previously been associated with dysregulated phenylalanine catabolism and cardiac impairment in mice (Czibik et al., 2021), and age-related reduction in creatinine clearance has been a key marker of decline in kidney function (Weinstein & Anderson, 2010). While these associations have generally been previously reported (Panyard et al., 2022), we confirmed their relationship with age in a multi-cohort setting. Furthermore, we found that metabolic associations with mortality well replicated previously reported findings (Deelen et al., 2019). We found that some of these metabolites were related to mortality in a direction consistent with the relationship with age including creatinine, phenylalanine and triglycerides, some age-related metabolites had neutral or non-significant relationship with mortality, while others particularly DHA, omega-3 fatty acids and the degree of unsaturation of fatty acids showed inverse relationships with mortality. Given that circulating metabolites have distinct physiological and regulatory functions, we speculate that some metabolites showing different directions of association to mortality and age may in fact be offering a protective/ adaptive or neutral response to the physiological ageing processes. For instance, DHA is thought to reduce oxidative stress and inflammation by modulating cyclooxygenase, lipoxygenase and cytochrome P450 lipid mediator activities (Swanson, Block, & Mousa, 2012; Zhang et al., 2013).

The models presented trained on CA include metabolites with neutral and potentially adaptive metabolic effects, yet remarkably still provide additional prediction of mortality, suggesting the models are capturing a higher-level picture of metabolic ageing, which overall contributes to mortality risk. While ageing markers trained on mortality are more sensitive to ageing risk factors and show improved prediction of age-related disease generally (Lu et al., 2019), they will to a greater extent capture extrinsic contributions, such as early effects of disease, to metabolic ageing. Within this study, we found that metabolic age models were sensitive to classical and modifiable risk factors of mortality (Stringhini et al., 2017), and also related to clinical biomarkers of system function, including blood pressure, C-creative protein, forced expiratory volume and glomerular filtration rate. Unexpectedly, heavy alcohol use appeared to be negatively associated with some metabolomic age models, which may be related to the effects alcohol consumption has on metabolites such as citrate (Wurtz et al., 2016), illustrating a limitation of the metabolic modelling approach for certain risk factors.

### Strengths and limitations

The use of multiple cohorts covering most of adult life is one of the strengths of this study and particularly important for analysis of metabolites, which may be impacted by both endogenous factors such as ageing and exogenous factors such as diet, since the relationship between age and exogenous factors (cohort effects) will likely be stronger within single cohorts. The use of some repeat samples, although in limited number, also increases the ability to detect endogenous ageing effects, while the use of fasting samples in our training set has lessened the possible influence of diet. The comparison cohort, UKB, is based on non-fasting samples, which can explain some of the relatively poor replication of the association with age in UKB. Another important limitation is that this study was mainly based on cross-sectional data, which is more susceptible to cohort effects than studies based on longitudinal data and does not allow assessment of trajectories of ageing over time (Ala-Korpela et al., 2023). Furthermore, the use of only Northern European cohorts may limit generalizability to other populations. Nevertheless, the main strength was the use of large, independent training and test sets, allowing completely unbiased assessment of model performance.

Metabolic profiling based on NMR provides both strengths and limitations for development of ageing metrics in a multi-cohort setting. The main advantage is that it is inherently quantitative, enabling comparable analyses of datasets across cohorts, and captures both small molecules and lipid metabolites. It is also high-throughput and cost-effective allowing the large population samples required for precise estimation of age-associations. The main limitation is the lower coverage of NMR compared to mass-spectrometry based methods, meaning that only the most abundant metabolites are detected, and many important and specific age-related metabolites may be missed. Identified metabolites such as citrate and DHA are undoubtedly important components of the ageing metabolome, being both among those most strongly associated with age in previous studies that employed broader MS-based analysis (Darst, Koscik, Hogan, Johnson, & Engelman, 2019; Menni et al., 2013). However, other key ageing metabolites such as steroids, acylcarnitine and tryptophan metabolites are not assayed by current NMR platforms. Future metabolic ageing studies will need to combine broad, highly sensitive metabolomics with careful control of technical variation to allow combination across studies.

### Conclusions

We have developed and tested various metabolic ageing metrics in a very large dataset. We found that the Deelen *et al*. model provides the most consistent prediction of mortality and all age-related diseases tested and is therefore a good candidate model for studies investigating metabolically mediated effects on lifespan. MARS, a non-linear method was found to improve prediction of CA over other modelling techniques. Our phenotypic ageing model directly predicts CA, while also providing good predictive ability of mortality and multiple age-related diseases and presents a good candidate for studies of overall metabolic ageing. Although we have shown that NMR metabolomics can only provide moderate prediction of CA across independent test sets, the technique provides valuable information regarding metabolic health, which is intricately linked to population ageing. We expect that future studies incorporating broader metabolomic analytical techniques will allow more comprehensive and specific assessment of metabolic ageing. These models may have utility for large-scale epidemiological analysis, allowing assessment of ageing risk factors and mechanisms and stratification and identification of at-risk groups.

## Acknowledgements

We are extremely grateful to all the families who took part in this study, the midwives for their help in recruiting them, and the whole ALSPAC team, which includes interviewers, computer and laboratory technicians, clerical workers, research scientists, volunteers, managers, receptionists and nurses. PBM acknowledges the support of the National Institute for Health and Care Research Barts Biomedical Research Centre (NIHR203330); a delivery partnership of Barts Health NHS Trust, Queen Mary University of London, St George’s University Hospitals NHS Foundation Trust and St George’s University of London.

## Authors’ contributions

OR conceived and designed the study. C-H.L performed most of the statistical analyses and MM performed the statistical analyses in UKB. C-H.L and OR drafted the manuscript. All other coauthors helped with data management, supervised data collection and curation, and managed the cohort studies. All authors have read and contributed to final drafting of the manuscript.

## Data availability statement

The data that support the findings of this study are available upon application to steering committee of each cohort.

## Abbreviations

NMR: nuclear magnetic resonance
MAE: mean absolute error
CI: confidence interval
FDR: false discovery rate
MARS: multivariate adaptive regression splines
VIF: variable inflation factor
VIP: variable importance of projection
CVD: cardiovascular disease
COPD: chronic obstructive pulmonary disease
CA: chronological age
BMI: Body Mass Index
ApoB: Apolipoprotein B
HDL: high-density lipoprotein
VLDL: very low-density lipoprotein
LDL: low-density lipoprotein
IDL: intermediate-density lipoprotein
TG: triglycerides
DHA: docosahexaenoic acid
SBP: systolic blood pressure
DBP: diastolic blood pressure
CRP: C-reactive protein
eGFR: estimated glomerular filtration rate
FEV1: forced expiratory volume in first second
UKB: UK Biobank
SABRE: Southhall and Brent Revisited
BWHHS: British Women’s Health Heart Study
UKCTOCS: The Collaborative UK Ovarian Cancer Screening Trial
WHII: Whitehall II
NSHD: MRC National Survey of Health and Development
YFS: Young Finns Study
NFBC1966: The 1966 Northern Finland Birth Cohort
CAPS: Caerphilly Prospective Study
ALSPAC: Avon Longitudinal Study of Parents and Children
UCLEB: UCL-LSHTM-Edinburgh-Bristol Consortium

## Supplementary Information

### Ageing models

#### 24 pruned metabolite variable set

Lactate, GlycA, Creatinine, Albumin, Tyr, Val, Phe, His, Gln, Ala, bOHbutyrate, Acetate, Glucose, Citrate, Omega_6, DHA, Sphingomyelins, HDL_TG, VLDL_size, S_HDL_FC, M_HDL_FC, XL_HDL_C, S_LDL_TG, XS_VLDL_FC

#### Study mortality score (mortality, standardised values)

S_LDL_TG × 0.01168 + Omega_6 × (- 0.10244) + Glucose × 0.00103 + His × (-0.07158) + Phe × 0.14083 + Val × (-0.01078) + GlycA × 0.07416

#### Deelen model (relative mortality, standardised values)

XXL_VLDL_L × log (0.80) + S_HDL_L × log (0.87) +VLDL_size × log (0.85) +PUFA_FA × log (0.78) +Glucose × log (1.16) +Lactate × log (1.06) +His × log (0.93) +Ile × log (1.23) +Leu × log (0.82) +Val × log (0.87) +Phe × log(1.13) +Acetoacetate × log(1.08) +Albumin × log(0.89) + GlycA × log(1.32)

#### Akker model (absolute values)

58.62 +

( ( ( Acetoacetate - 0.04319 ) / 0.03465 ) × 1.056 ) +

( ( ( Acetate - 0.0445 ) / 0.01919 ) × 0.6887 ) +

( ( ( Ala - 0.3006 ) / 0.0767 ) × -0.3769 ) +

( ( ( Albumin - 0.08774) / 0.006863 ) × -2.843 ) +

( ( ( APOA1 - 1.594 ) / 0.2002 ) × 0.9177 ) +

( ( ( APOB - 0.9732 ) / 0.2213 ) × 8.111 ) +

( ( ( Citrate - 0.09357 ) / 0.02787 ) × 2.501 ) +

( ( ( Creatinine - 0.07223 ) / 0.01835 ) × 2.168 ) +

( ( ( DHA - 0.1447 ) / 0.05478 ) × -0.8051 ) +

( ( ( Omega_3 - 0.4114 ) / 0.134 ) × 7.364 ) +

( ( ( FAW3_FA - 3.576 ) / 0.9525 ) × 2.75 ) +

( ( ( Omega_6 - 3.871 ) / 0.7695 ) × 63.88 ) +

( ( ( FAW6_FA - 33.76 ) / 3.567 ) × -10.21 ) +

( ( ( Glucose - 4.8 ) / 1.611 ) × 1.394 ) +

( ( ( Gln - 0.4528 ) / 0.07966 ) × 3.844 ) +

( ( ( GlycA - 1.359 ) / 0.2039 ) × 0.1866 ) +

( ( ( HDL2_C - 0.8999 ) / 0.3046 ) × -161.3 ) +

( ( ( HDL3_C - 0.4698 ) / 0.06615 ) × -35.57 ) +

( ( ( HDL_C - 1.37 ) / 0.3277 ) × 187.4 ) +

( ( ( HDL_size- 9.972 ) / 0.2498 ) × 1.254 ) +

( ( ( His - 0.05925 ) / 0.01482 ) × -2.084 ) +

( ( ( IDL_C - 0.6855 ) / 0.1948 ) × -0.04409 ) +

( ( ( IDL_L - 1.069 ) / 0.2802 ) × -3.969 ) +

( ( ( Ile - 0.05285 ) / 0.02023 ) × -1.844 ) +

( ( ( L_LDL_L - 1.171 ) / 0.3441 ) × -23 ) +

( ( ( LA - 3.099 ) / 0.6966 ) × -3.273 ) +

( ( ( Lactate - 1.232 ) / 1.032 ) × 1.868 ) +

( ( ( LDL_C - 1.488 ) / 0.5062 ) × 15.22 ) +

( ( ( LDL_size - 23.65 ) / 0.119 ) × 0.2465 ) +

( ( ( Leu - 0.06167 ) / 0.01617 ) × -4.118 ) +

( ( ( M_HDL_L - 0.8082 ) / 0.1624 ) × -5.544 ) +

( ( ( M_LDL_L - 0.6606 ) / 0.2066 ) × 33.28 ) +

( ( ( M_VLDL_L - 0.6664 ) / 0.3765 ) × -6.233 ) +

( ( ( MUFA - 2.904 ) / 0.9051 ) × -8.85 ) +

( ( ( MUFA_FA - 24.83 ) / 3.645 ) × -68.24 ) +

( ( ( Phosphatidylc - 1.985 ) / 0.3749 ) × -3.59 ) +

( ( ( Phe - 0.04339 ) / 0.008491 ) × 2.939 ) +

( ( ( PUFA - 4.282 ) / 0.8429 ) × -67.3 ) +

( ( ( PUFA_FA - 37.34 ) / 3.682 ) × -61.45 ) +

( ( ( S_HDL_L - 1.004 ) / 0.1017 ) × 4.017 ) +

( ( ( S_LDL_L - 0.4282 ) / 0.1244 ) × -10.42 ) +

( ( ( S_VLDL_L - 0.7053 ) / 0.2318 ) × -9.983 ) +

( ( ( Total_C - 4.42 ) / 0.9903 ) × -28.22 ) +

( ( ( Total_TG- 1.397 ) / 0.6807 ) × 4.872 ) +

( ( ( SFA - 4.375 ) / 0.9667 ) × -19.06 ) +

( ( ( SFA_FA - 37.84 ) / 1.867 ) × -31.47 ) +

( ( ( Sphingomyelins - 0.4548 ) / 0.08845 ) × 1.705 ) +

( ( ( Cholines - 2.293 ) / 0.3814 ) × -3.358 ) +

( ( ( Total_FA - 11.56 ) / 2.482 ) × 23.67 ) +

( ( ( Phosphoglyc - 1.898 ) / 0.3669 ) × 5.78 ) +

( ( ( Tyr - 0.06123 ) / 0.01556 ) × 2.209 ) +

( ( ( UNSAT - 1.215 ) / 0.0745 ) × -0.49 ) +

( ( ( Val - 0.1574 ) / 0.03815 ) × 1.656 ) +

( ( ( VLDL_C - 0.8767 ) / 0.2799 ) × 15.42 ) +

( ( ( VLDL_size- 36.79 ) / 1.361 ) × 3.839 ) +

( ( ( XS_VLDL_L - 0.5663 ) / 0.1263 ) × 5.976 )

### Cohort information

#### Avon Longitudinal Study of Parents and Children

The Avon Longitudinal Study of Children and Parents (ALSPAC) was established to understand how genetic and environmental characteristics influence health and development in parents and children. All pregnant women resident in Avon, UK with expected dates of delivery 1st April 1991 to 31st December 1992 were invited to take part in the study. Of the original 14,541 initial pregnancies, 338 were from a woman who had already enrolled with a previous pregnancy, meaning 14,203 unique mothers were initially enrolled in the study. As a result of the additional phases of recruitment, a further 630 women who did not enroll originally have provided data since their child was 7 years of age. This provides a total of 14,833 unique women (G0 mothers) enrolled in ALSPAC as of September 2021. G0 partners were invited to complete questionnaires by the mothers at the start of the study and they were not formally enrolled at that time. 12,113 G0 partners have been in contact with the study by providing data and/or formally enrolling when this started in 2010. 3,807 G0 partners are currently enrolled. Study data were collected and managed using REDCap electronic data capture tools hosted at the University of Bristol (Harris et al., 2009). REDCap (Research Electronic Data Capture) is a secure, web-based software platform designed to support data capture for research studies. Consent for biological samples has been collected in accordance with the Human Tissue Act (2004) and ethical approval for the ALSPAC study was obtained from the ALSPAC Ethics and Law Committee and the Local Research Ethics Committees. Informed consent for the use of data collected via questionnaires and clinics was obtained from participants following the recommendations of the ALSPAC Ethics and Law Committee at the time. Please note that the study website contains details of all the data that is available through a fully searchable data dictionary and variable search tool: http://www.bristol.ac.uk/alspac/researchers/our-data/

#### Northern Finnish Birth Cohort 1966

The study was started in the two Northernmost provinces in Finland (Oulu and Lapland) in the year 1965 when the mothers were pregnant. Data on the individuals born into this cohort was collected since the 16th gestational week as well as their mothers and, to a lesser extent, fathers. The cohort included 12,055 mothers and they had 12,068 deliveries (13 women delivered twice). Cases belonging to survey were determined by the calculated term. A small percentage of the births occurred towards the end of 1965 and early in 1967. The calculated term, as was customary at that time, was counted from the first day of the last menstrual period. Where this date was unknown the expected term was estimated from the date of commencement of foetal movements and progress of the pregnancy. The study covered all live born and stillborn infants with birth weight of 600 grams or more. According to the Finland’s central Office of Statistics, births in the study area during 1966 totaled 12,527, so study population comprised 96.3% of all births during 1966 in that area. Altogether 12,231 children were born into the cohort, 12,058 of them live-born. The original data have been supplemented by data collected with postal questionnaires at the ages of 1, 14, 31 and 48 years and various hospital records and national register data.

#### Young Finns Study

The program was launched in Finland in the late 1970’s to study cardiovascular risk in the youth. The multi-centre study, called The Cardiovascular Risk in Young Finns, was designed to study the risk factors and precursors of cardiovascular diseases and their determinants in children and adolescents. Two pilot studies were carried out in 1978 and 1979, and the first cross-sectional study in 1980. Thereafter, this cohort has been followed-up several times, and the latest field study was conducted in 2011/12. The first cross-sectional survey was conducted in 1980. Total sample size was 4,320 boys and girls in 6 age cohorts (aged 3, 6, 9, 12, 15 and 18). These subjects were randomly chosen from the national register. A total of 3,596 subjects (83.2% of those invited) participated in the study in 1980. After that, several follow-up studies of this cohort have been conducted. The participation rates in the follow-up studies have varied between 60 and 80%. In the latest follow-up in 2011/12 a total of 2,063 subjects were examined (57% of the original cohort).

#### MRC National Survey of Health and Development

The NSHD has informed UK health care, education and social policy for more than 50 years and is the oldest and longest running of the British birth cohort studies. Today, with study members in their seventies, the NSHD offers a unique opportunity to explore the long-term biological and social processes of ageing and how ageing is affected by factors acting across the whole of life. From an initial maternity survey of 13,687 of all births recorded in England, Scotland and Wales during one week of March 1946, a socially stratified sample of 5,362 singleton babies born to married parents was selected for follow-up. This sample comprises the NSHD cohort and participants have been studied 24 times. During their childhood, the main aim of the NSHD was to investigate how the environment at home and at school affected physical and mental development and educational attainment. During adulthood, the main aim was to investigate how childhood health and development and lifetime social circumstances affected their adult health and function and how these change with age. Now, as participants pass retirement age, the research team is developing the NSHD into a life course study of ageing.

#### Southall And Brent REvisited Study

The Southall And Brent REvisited Study (SABRE) is the largest tri-ethnic population-based cohort in the UK, involving nearly 5,000 European, Indian Asian and African Caribbean men and women. It investigates the causes of diabetes and disorders of the heart and circulation. The participants were aged 40-69 when first studied between 1989 and 1991. In 2008 – 2011 a comprehensive combined morbidity and mortality follow up was carried out, together with non-invasive clinical measurements in order to quantify sub-clinical disease. SABRE visit 2 tested hypotheses generated from the Southall and Brent baseline studies and ongoing mortality follow-up. SABRE Visit 3 (25-year follow-up visit) started in July 2014 and will collect data on participants and their partners. The aims of the study are to build on what has been learned from the first study. Changes in the health of the heart and circulation will be measured, with a special focus on the health of the blood vessels of the brain, as well as early signs of diabetes. The study will also look at physical function and how well (or otherwise) people are keeping as they get older.

#### Whitehall II Study

The Whitehall II study was established to investigate the causes of social inequalities in health (Marmot & Brunner, 2005). A cohort of 10,308 participants aged 35-55, of whom 3,413 were women and 6,895 men, was recruited from the British Civil Service in 1985. Since this first wave of data collection, self-completion questionnaires and clinical data have been collected from the cohort every two to five years with a high level of participation. The Whitehall II study has shown the importance of psychosocial factors such as work stress and work-family conflict in heart disease and diabetes. These are in addition to the contribution of unhealthy behaviours and traditional risk factors (such as high blood pressure).

#### Caerphilly Prospective Study

The Caerphilly Prospective Study (CAPS) was set up by the MRC Epidemiology Unit (South Wales). At that time, it was the fifth prospective study of cardiovascular disease in the United Kingdom, although only the second population-based study, after the British Regional Heart Study. Its initial aims were to examine the importance of lipids, haemostatic factors, and hormones such as testosterone, cortisol and insulin (Lichtenstein et al 1987) in the development of ischaemic heart disease (IHD). Subsequently, other hypotheses were included with a specific interest in platelet function, and psychosocial variables. With the ageing of the cohort, additional outcomes have been included in particular stroke, hearing problems and cognitive function. The initial design attempted to contact all men aged 45 to 59 years from the town of Caerphilly and adjoining villages. Two thousand, five hundred and twelve subjects (response rate 89%) identified from the electoral register and general practice lists were examined between July 1979 until September 1983 (phase I). Men were initially seen at an evening clinic, where they completed a questionnaire, had anthropometric measures and an ECG taken. They also completed a food frequency questionnaire at home (Fehily et al 1994). They subsequently re-attended an early morning clinic to have fasting blood samples for a wide variety of tests. Quality control was examined by the use of both "blind" split samples as well as a second repeat measure on a random sub-sample to examine intra-individual variation.

#### UK Collaborative Trial of Ovarian Cancer Screening Longitudinal Women’s Cohort

The cohort is the bioresource built in the course of the United Kingdom Collaborative Trial of Ovarian Cancer Screening (UKCTOCS) (Jacobs et al., 2016). The latter is designed to test the hypothesis that ovarian cancer screening can save lives by detecting the disease earlier. Between April 2001-Sept 2005, 202,638 postmenopausal women, aged 50-74 years were recruited through 13 trial centers in England, Wales and Northern Ireland. Women were randomly allocated to one of three groups (i) control (C) - no screening (ii) multimodal screening (MMS) - annual blood test for serum CA125 measurement. The results were interpreted using the ‘Risk of Ovarian Cancer Algorithm’, with transvaginal ultrasound as a second line test in case of abnormality (iii) ultrasound screening (USS) – annual and second line tests were transvaginal scans. Women in the screen arms underwent a total of 673,765 annual screens till 31st December 2011. The whole cohort is linked to multiple UK electronic health records with ongoing active follow-up.

#### British Women’s Heart and Health Study

The British Women’s Heart and Health Study (BWHHS) is a prospective cohort study of cardiovascular disease in women aged over 60 years, in England, Scotland and Wales (Lawlor, Bedford, Taylor, & Ebrahim, 2003). Set up in 1999 to complement the British Regional Heart Study (BRHS), to describe and establish risk factors and the differences in their impact in women compared to the men followed up by the BRHS. The study selected women at random from 24 GP practices, in 23 towns from 1999 to 2000. Of the 7,296 invited, 4,286 (60%) were recruited and attended the baseline examinations and completed questionnaires. Follow-up consisted of postal questionnaires and regular reviews of General Practitioners’ medical records.

#### UK Biobank

UK Biobank (UKB) is a very large and detailed prospective study with over 500,000 UK participants, collected and continues to collect extensive phenotypic and genotypic detail about its participants, including data from questionnaires, physical measures, sample assays, accelerometry, multimodal imaging, genome-wide genotyping and longitudinal follow-up for a wide range of health-related outcomes. Participants were 40–69 years old when recruited in 2006–2010. UK Biobank is available for open access, without the need for collaboration, to any bona fide researcher who wishes to use it to conduct health-related research for the benefit of the public (Sudlow et al., 2015). Incident cases of CVD, COPD and dementia were accessible through algorithms made publicly available by the UKB using data from hospital and death registers: https://biobank.ndph.ox.ac.uk/showcase/showcase/docs/alg_outcome_stroke.pdf, http://biobank.ndph.ox.ac.uk/showcase/showcase/docs/alg_outcome_mi.pdf, http://biobank.ndph.ox.ac.uk/showcase/showcase/docs/alg_outcome_dementia.pdf, http://biobank.ndph.ox.ac.uk/showcase/showcase/docs/alg_outcome_copd.pdf. For T2DM incident cases we used the following International Classification of Diseases 10^th^ revision (ICD-10) codes: E11.0, E11.1, E11.2, E11.3, E11.4, E11.5, E11.6, E11.7, E11.8, E11.9, while incident diagnosis of cancer recorded as all cancer (ICD-10: C00-97) including melanoma.

## Supplemental Figures

**Supplemental Figure 1.**
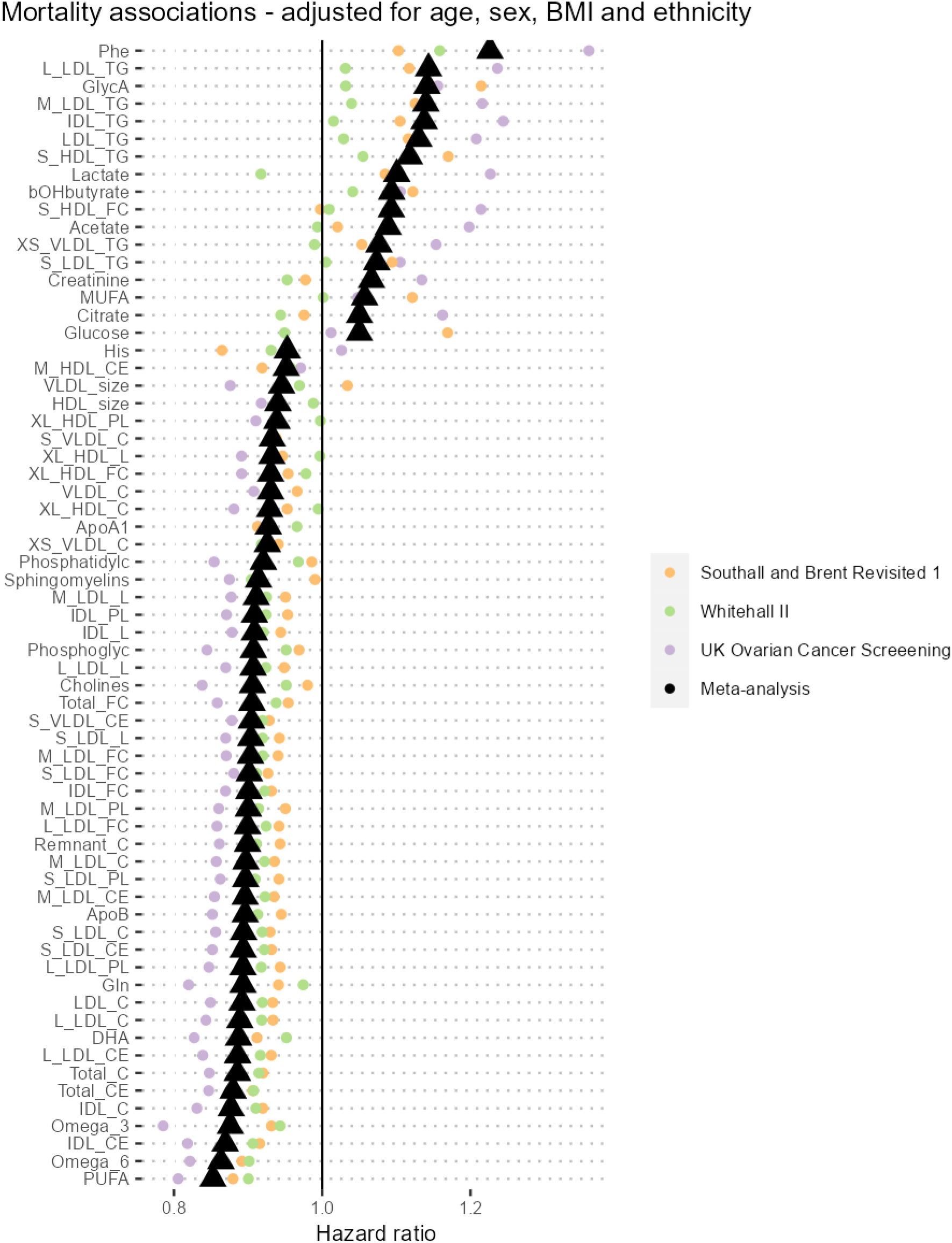
NMR metabolome associations with all-cause mortality in WHII, SABRE, and UKCTOCS. Cohort-specific Cox proportional hazards regression models were adjusted for age, sex, BMI and ethnicity; fixed effected meta-analysis was performed to pool together individual cohort estimates, and significant positive and negative associations with mortality after correcting for false discovery rate (q < 0.05) were shown.

**Supplemental Figure 2.**
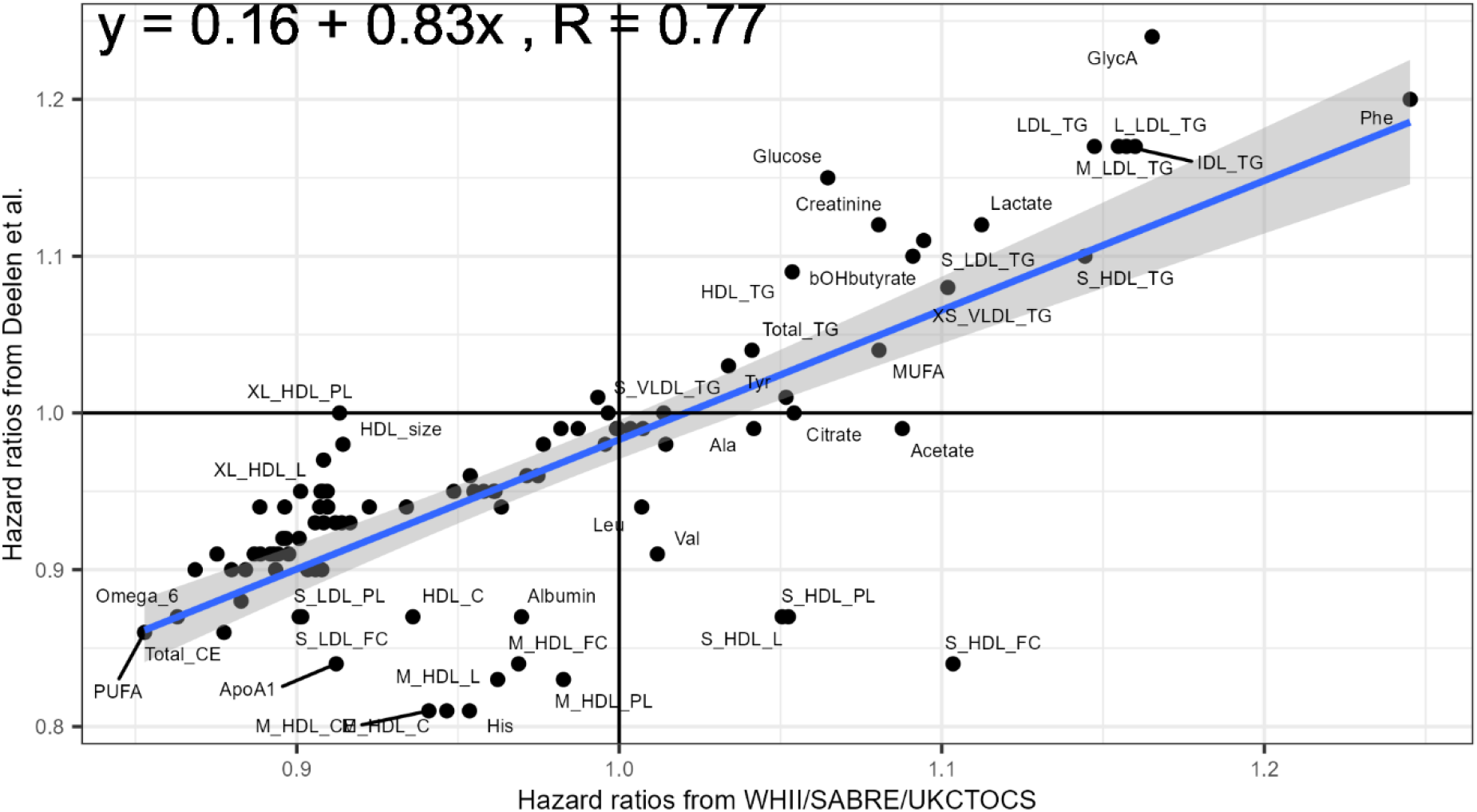
Mortality associations compared to published values obtained from Deelen’s et.al. Mortality coefficients values were obtained from Deelen’s published paper (Deelen et al., 2019). “Unadjusted model coefficients” were extracted from Deelen’s paper Supp Data 1. Deelen’s mortality coefficients were adjusted for age and sex and cohort. The estimates represent ln(hazard) per standard deviation change in metabolite level.

**Supplemental Figure 3.**
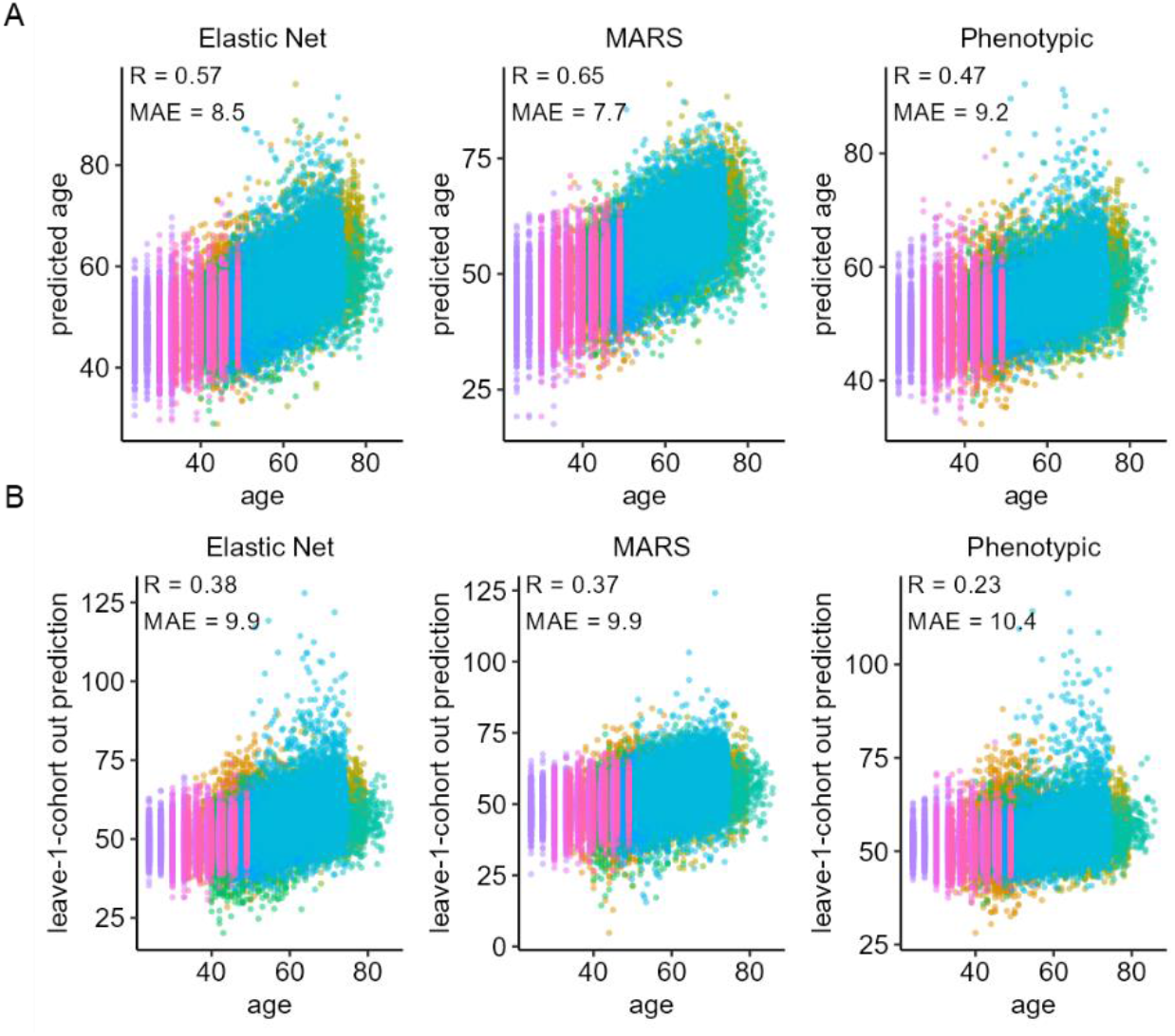
Fitted model prediction (top) and leave-one-cohort-out cross validation of modelled age predictions (bottom).

**Supplemental Figure 4.**
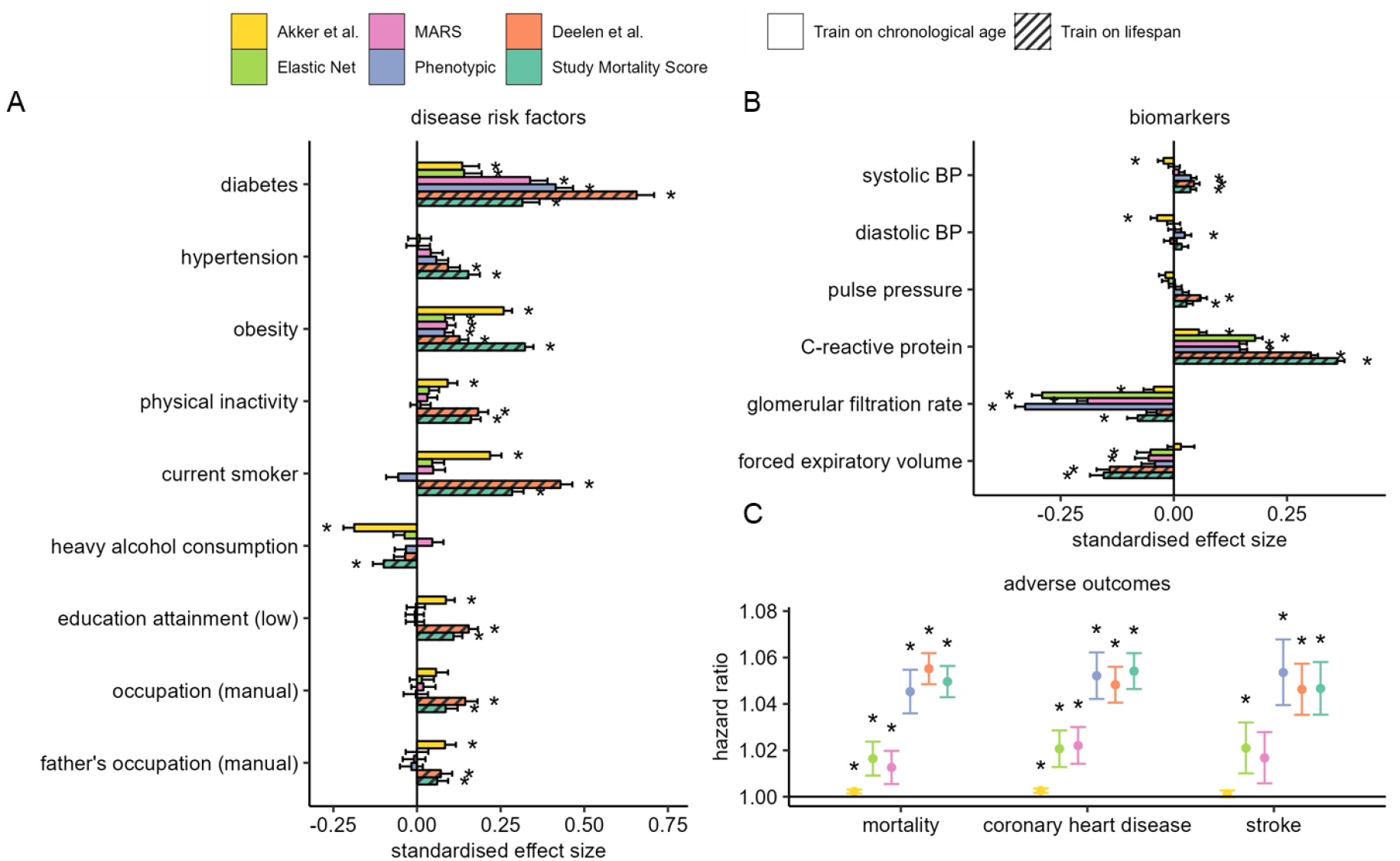
Model associations in UCLEB additionally adjusted for BMI A) Associations with known disease risk factors. Estimates represent standard deviation (SD) change in metabolomic age associated with exposure which have been categorised into <u>binary</u> variables. B) Associations with age-related biomarkers. Estimates represent standard deviation change in metabolomic age associated with 1 SD unit change in biomarker levels. To avoid individuals from being accounted for more than once in the analysis, samples from YFS2001 and YFS2007, NFBC1966 (31y), and SABRE2 were excluded in the disease risk factor analysis, and subsequently up to 28,000 samples were included. C) Associations of metabolomic age models with adverse incident health events. Cox proportional regression models were adjusted for chronological age, sex, BMI and ethnicity.

